# Mapping smartphone-based eye-tracking behavior across Japanese individuals on the pareidolia test

**DOI:** 10.1101/2024.06.20.24308648

**Authors:** Gajanan S. Revankar, Kota Furuya, Etsuro Mori, Maki Suzuki, Yoshiyuki Nishio, Issei Ogasawara, Yuki Yamamoto, Abhay M. Aradhya, Abhishek C. Salian, Varad V. Kajarekar, Ashwin M. Jagadeesh, Seema S. Revankar, Arya A. Revankar, Natsuki Yoshida, Chizu Saeki, Tatsuhiko Ozono, Daisaku Nakatani, Hideki Mochizuki, Manabu Ikeda, Ken Nakata

## Abstract

Pareidolias are illusionary phenomena wherein ambiguous forms appear meaningful. In clinical research, pareidolias have been studied using paper or desktop test formats to deconstruct visuo-perceptual mechanisms. Translating this work on to an accessible, scalable setup such as smartphones is currently unknown. Here, we designed a smartphone-based pareidolia test to study visual processes affecting gaze behavior of cognitively healthy individuals using a standard, native front-facing camera. We optimized our system using machine learning and explored the challenges involved in user behavior, demographic specificity, and test functionality. We performed our experiments on 52 healthy Japanese adults, aged between 50 to 80 years who underwent MMSE and the smartphone test for pareidolias. Gaze movements on the 15-min, user-centric evaluation was calibrated to every individual. Results showed test responses with minimal differences with respect to age, sex, and completion time. Personalized calibrations improved the model’s prediction performance and quantification of gaze tracking metrics aligned with that of commercial grade eye-trackers. Our findings demonstrate the applicability and scalability of pareidolia testing on smartphone platforms.

## Introduction

Gaze-tracking has been extensively employed in both research and the development of solutions across medical and neuroscientific areas^1–3^. Field-based studies for gaze capture utilize specialized, expensive hardware, although the gap is rapidly being addressed by several scalable desktop and mobile solutions^4,5^. Recent advances in machine learning (ML) has allowed implementation of eye-tracking (ET) on smartphone-based systems providing a stable framework for usage across vision research and healthcare^5–7^.

The predominant usage of ET in neuroscience has been to deconstruct attentional and visuo-perceptual mechanisms^2,8^. In our prior work, we used ET (with synchronized EEG capture) to study a visuo-perceptual phenomenon called pareidolia - illusionary events wherein uncertain shapes or forms appear meaningful^9,10^. Performed in a clinical setting, we demonstrated the use of ET-EEG to deconstruct neural correlates that affect frontal cortex and attentional dynamics in Parkinson’s disease patients^11^. We employed a commercial grade ET system equipped with multiple high spatio-temporal resolution infrared cameras to monitor user responses as they viewed images presented on a standard 24” desktop monitor. Though its usage in a research context may be justified, deployment of such hardware/software system poses significant difficulties in resource-constrained environments such as tertiary-care centers or smaller clinics.

Pareidolias hold considerable clinical significance. While pareidolias may seem inconsequential in healthy individuals, they serve as crucial indicators for neuropsychiatric disorders. Clinically, with the loss of insight, pareidolias act as early behavioral markers for psychosis in Alzheimer’s disease (AD)^12,13^, Dementia with Lewy body disease (DLB)^14,15^ or Parkinson’s disease (PD)^16–18^, establishing it as a significant prognostic indicator for disease progression^15^. However, individual variations of pareidolia is common among cognitively normal adults and such perceptual tendencies are also known to vary across cultures requiring some degree of demographic personalization^19,20^.

The objective of our current study was to study the visuo-perceptual phenomenon of pareidolia translated from a desktop-ET environment on to a smartphone for a known demographic. To that end, we developed an in-house ET software that uses existing smartphone front-facing camera for gaze tracking. Using this ET software, we outlined the eye-movement behavior across cognitively normal Japanese individuals on a standardized visuo-perceptual test called the Noise pareidolia test (NPT)^21^. The NPT is a neuropsychological test to evaluate and quantify face-related misperceptions or face-pareidolias^22^. We also explored the implementational challenges associated with developing smartphone ET in terms of usage, accuracy of measurements and limited computational capacity.

Within this scope, we outline our findings regarding the usage and behavioral characteristics observed during the pareidolia test among healthy Japanese participants. Additionally, we delve into the gaze tracking methodology employed, which lays the groundwork for the development of a digital pareidolia test on a smartphone.

## Methods

### General information and consent

52 Healthy participants (26F, 26M) were enrolled in this cross-sectional study. Participants were recruited via a large-scale online survey across Osaka prefecture, Japan for participation in the study. Individuals were between 50 to 80 years of age, without any past or present neurological problems. Participants were tested with a near-vision Snellen chart integrated in the smartphone to include normal or corrected-to-normal vision subjects only.

Each individual provided their explicit and written informed consent to data collection that informed them about collecting the front-facing camera feed for research analyses purposes, and the potential risks involved in performing gaze tasks for several minutes (e.g., eye strain, fatigue). Monetary compensation was provided, and all participants retained the option to have their data deleted at any time. The smartphone app was designed to be around 15 min in length and was conducted in a controlled lab setting. The Osaka University institution review board cleared the protocol for the study to be performed in the Department of Neurology, Osaka University, Japan in accordance with the ethical standards of the Declaration of Helsinki (IRB Approval number – 22307).

### Experiment flow

Following informed-consent, participants underwent i) Mini-mental State Examination, Japanese version (MMSE-J)^23^ and ii) the digital Noise pareidolia test (dNPT). MMSE was administered by a clinical psychologist or a neurologist. The dNPT is a digital version of the paper-based NPT which comprises of 40 black and white images, with a face embedded in 8 of the 40 images. Participants must locate and identify faces accurately, and any misidentification of noisy areas as faces are marked as pareidolia. As opposed to the paper-test, images in dNPT were randomized to prevent a learning effect^24^.

Participants performed the dNPT test on an Android smartphone [Samsung A32 with screen dimensions 164mm (h) x 76mm (w), portrait mode, 13MP front camera], that was fixed on a smartphone mount, with participants sitting at approx. 30 cm from the screen. Screen resolution and screen brightness were set at a fixed level across all participants. The rooms were lit using standard multiple fluorescent light sources, with a luminosity range of approximately 300 to 500 lux. We took care to position participants in a way that avoided creating glares, reflections, or shadows on the testing apparatus (Supplementary Figure-1A). Appropriate brightness range on the face was ensured so that face and eyes were clearly visible. Brightness values for face were set between 50 – 89 pixels intensity (brightness levels calculated as per RGB pixel values). For eye-movement calibration, participants were asked to fixate on a pulsating black circular dot stimulus that appeared on a light-grey background with sufficient contrast. The dot, approx. 1cm in diameter, moved slowly from top-left to the bottom-right in a zig-zag fashion for 150 s. This calibration sequence was considerably longer in duration than prior studies but allowed a significant buffer to deal with lost frames during calibration. Images from the 13-megapixel front-facing camera were recorded at 1080p resolution at 30 Hz with timestamps synchronized with the marker location.

### Model details

For the base model training, MIT GazeCapture dataset was used since no gaze models specific to Japanese populations were available. The open-source MIT GazeCapture dataset includes data capture from mobile and tablet devices. Data diversity was quite extensive across various attributes such as sample size (Total N= 1474, with smartphone = 1249 participants, with tablet device = 225 participants), age groups (young to older adults), the use of spectacles, varying lighting conditions (indoors / outdoors), etc. For our analysis, we focused solely on smartphone data captured in portrait orientation. Post-processing was performed to remove bad samples. The base model that was defined from this dataset was approx. 80% of the total samples in the dataset (Total samples = 819,427; Final sample size = 659,930).

We used a multi-layer feed-forward Convolutional Neural Network (CNN). Compared to traditional Machine Learning models like Support Vector Regressor, etc, CNN captures complex features and composition of features automatically and has strong predictive abilities. The base CNN model was trained first on MIT GazeCapture dataset and then fine-tuned on a subset of the Japanese population (N=52 participants in this study). Feature-engineering techniques were used to process data for more meaningful features to personalize the model specific to Japanese face and eye capture data. The model architecture was designed to be lightweight and computationally less expensive. Model compression was conducted to further decrease the parameter count and ensure compatibility with smartphones. During fine-tuning, all the layer weights of the pre-trained base model were allowed to be updated until the model converged.

Eye regions were cropped based on the eye corner landmarks, scaled to 128 × 128 × 3 pixels, and fed through two identical convolutional layer blocks. Other features like eye corners and face area were fed through fully connected layers blocks to ultimately combine and get the gaze location on screen. We used average pooling and ReLU activation functions in the model. Description of model architecture is shown in Supplementary information (Supplementary Figure-2).

### Data Collection and Statistical analysis

Data was collected through an android app operating on OS version 13. While the app was functional from OS versions 11 onwards, formal testing on versions older than v13 was not conducted. For eye-movement analysis across the stimuli images, the methodological principles were as defined in our prior research^9^. Briefly, the front-camera video records participants’ gaze for each of 40 stimuli images on the smartphone screen. Areas of interest for faces and for noise patches were predefined on the software using a grid-based system. Fixations were defined as any 30 frames of continuous participant gaze within the same region, indicating a single fixation. 30 frames meant 30 continuous predictions (gaze locations) were within 50 pixels of distance for every continuous pair of predictions. This threshold was set after a trial-and-error estimation of the optimal count required to plot the visual tracking geometry. Saccades were defined as distances between 2 successive fixations that exceeded this threshold. Together, these metrics were used to calculate a heat-map index i.e. fixation and saccade counts, for comparing the visual tracking for target (faces) and non-target (noise) stimuli.

Demographic and MMSE data were stored in paper-based case report forms. Digital data from dNPT were saved on a database in a cloud server and the processing and readouts were done offline. Statistical analysis was performed on JASP (version 0.18.3). Outcome variables included pareidolia scores (false-positives), missed responses (false-negatives) and correct responses (true-positives and true-negatives) for dNPT. To report significant differences between groups, the statistical value was set to p < 0.05.

Due to the nature of NPT, pareidolia scores are almost always positively skewed distributions^9,24^. For 2-group comparison, Welch’s t-test or Mann Whitney-U test were performed depending on satisfactory assumptions of normality. For correlational analysis, Pearson’s r coefficient was reported for continuous data, and Chi-squared tests for binary/categorical variables.

### Data and Code Availability Statement

The data analyzed in this study will be provided by the corresponding author upon request, subject to reasonable conditions. The paper version of NPT is accessible under an open license for research purposes. The software code utilized in this study relies on internal tooling and infrastructure and is protected by patent (application number: JP2022-179766). Gaze estimation was done using PyTorch (v2.1.0) running on Python 3.10. Image preprocessing was performed using mediapipe v0.10.0, and gaze heatmaps were generated using heatmappy library (a Python library).

## Results

### General overview

52 participants (26F and 26M) were enrolled, aged 63.8 years ±8.1 (Mean, SD) with an MMSE score of 29.0 ±1.1. Notably, 2 participants had an MMSE score of 25. Participants took approx. 15 minutes to complete the full experiment (14.9 min ±2.7), which included onboarding via instructions, calibrations, some practice test images and finally the main test – dNPT, which took approx. 6 min (5.6 min ±1.6). Older participants took longer to complete the dNPT (Pearson’s r=0.27, p=0.053, Figure-2), and the completion time did not vary significantly across participants (Interquartile range: 4.7min to 6.4min). However, women were slightly quicker in terms of test completion (F: 5.4min ±1.6 vs M: 5.9min ±1.6).

**Figure-1:**
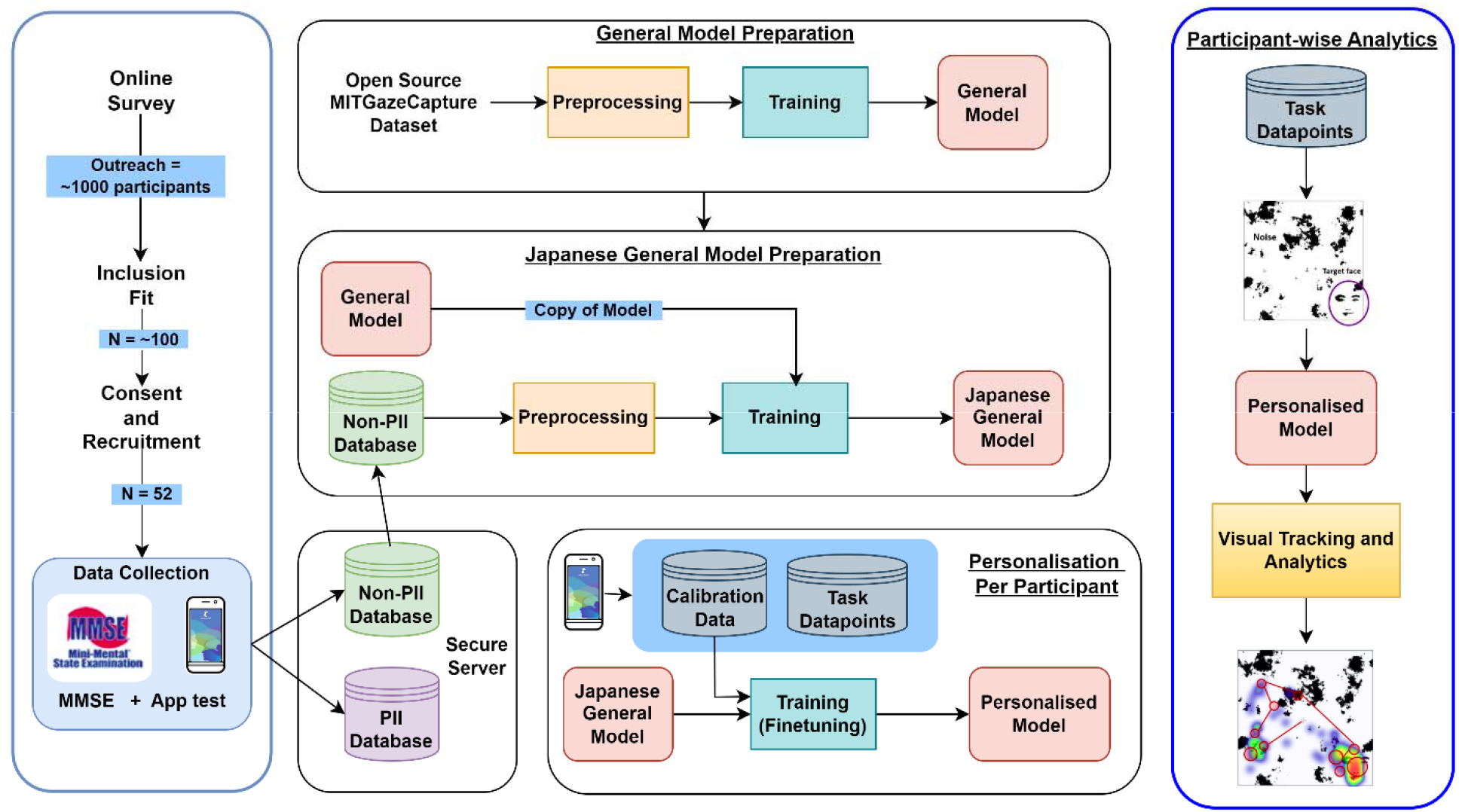
Overview of the evaluation protocol and development. Flowchart shows the recruitment of participants, model preparation and user analytics for eye-movement behavior. Legend: MMSE = Mini mental state examination, PII = Personal identifiable information.

**Figure-2:**
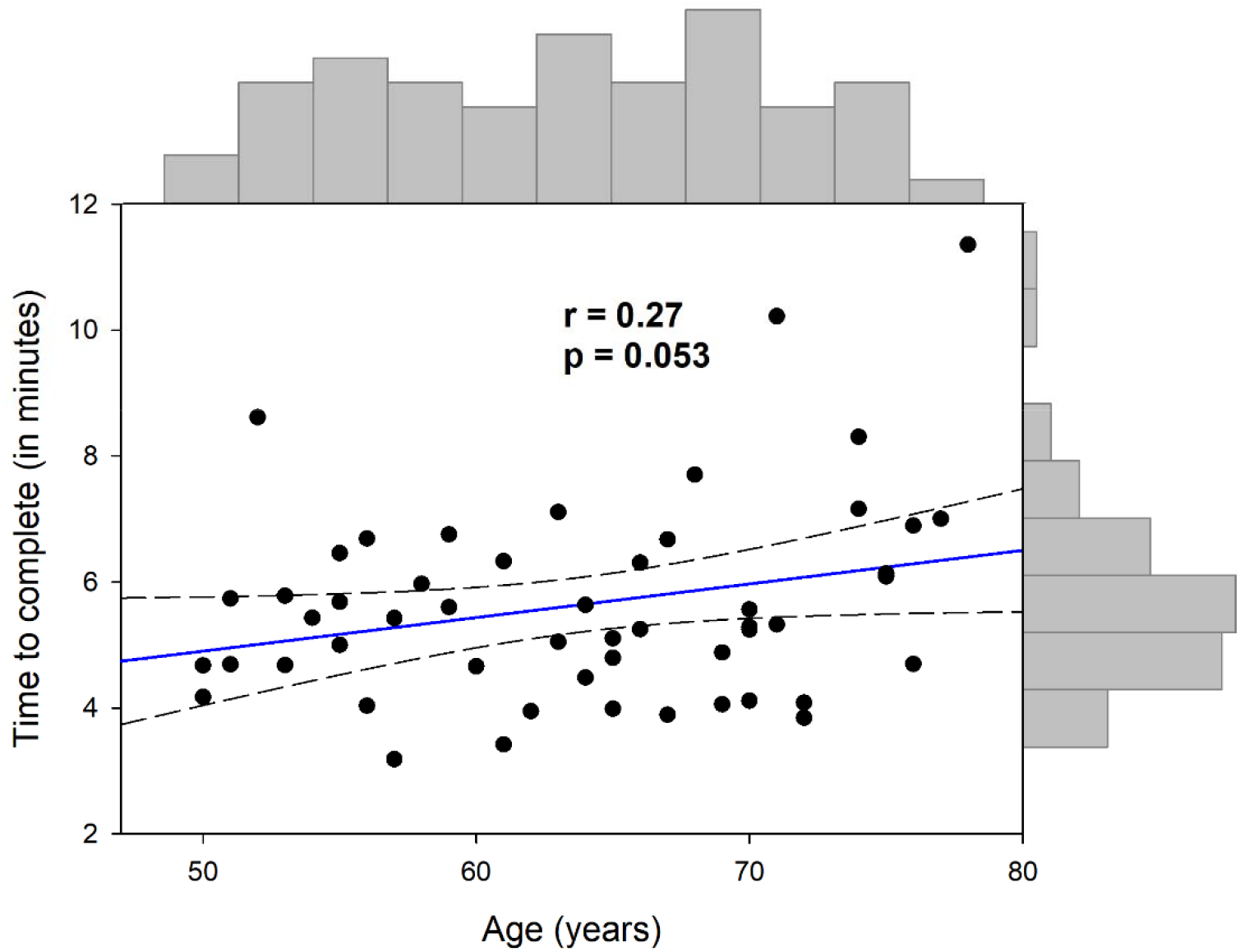
Correlation plot of completion time with respect to age.

Figure-2 shows correlation and distribution (outer axes) of the participants’ age with respect to the time to complete the core stimuli experiment. The regression line is shown in solid blue, with confidence intervals (CI) in dashed gray. Pearson’s r and p values are defined within the graphs.

Our testing was conducted in a well-lit environment with no frames being discarded based on uneven brightness levels (Mean environment brightness levels =137.76 ±14.00 SD; min=87.36 and max=181.29; pixel intensity ranging from 0 to 255). We observed a center bias with most participants positioned slightly to the left of the screen (left vs right = 92% vs 8%). However, handedness was not formally documented.

### Personalization for Japanese dataset

For gaze analysis, we obtained a total of 2068 samples and following postprocessing, trained our ET model on 1934 valid samples. 32 of 52 participants wore eyeglasses. We fine-tuned our Japanese dataset with respect to eye-shape, eyelid inter-palpebral fissure size (the distance between upper and lower eyelid), and presence of eyeglasses (Supplementary Figure-1B). Inclusion of additional variables/features did not seem to affect the validation and training losses on the model numbers. Reducing the eye-crop boundaries from the base-model to Japanese eye-crop data worsened the calibration error rates for our model (Mean error from 1.40 cm to 2.14 cm). We therefore carried out a default eye cropping method with predefined landmarks on the participants’ faces.

### Calibration accuracy

A 50-point grid was used for calibration. 15 (of the 52) participants (7F, 8M; 5 with eyeglasses) completed the full 50-point calibration sequence, 30 participants had a median calibration completion of 38-points (1^st^ Quartile, 3^rd^ Quartile = 31, 42), and 7 participants had poor tracking quality with calibration of less than 25-points (Median = 21, 1^st^ Quartile, 3^rd^ Quartile = 20, 22). We observed that the prediction errors were high for margins / corner areas when compared to the central screen areas on the smartphone (Supplementary Figure-3).

To get the best possible estimate of the baseline prediction errors across all 50-points, we included only those participants that completed the calibration sequence in its entirety. Accordingly, the base model error for our cohort was 1.40 cm ±0.51 (Mean, SD), and the personalization (finetuning or retraining) of model resulted in a 62.7% reduction in error equaling 0.52 cm ±0.15. Calibration error rates for the best and the worst participant were 0.31 cm ±0.16, and 0.91 cm ±0.81 error respectively (5^th^, 95^th^ percentile = 0.34cm, 0.81cm). At a viewing distance of approx. 30 cm. the error plots across the smartphone screen areas are shown in Figure-3.

**Figure-3:**
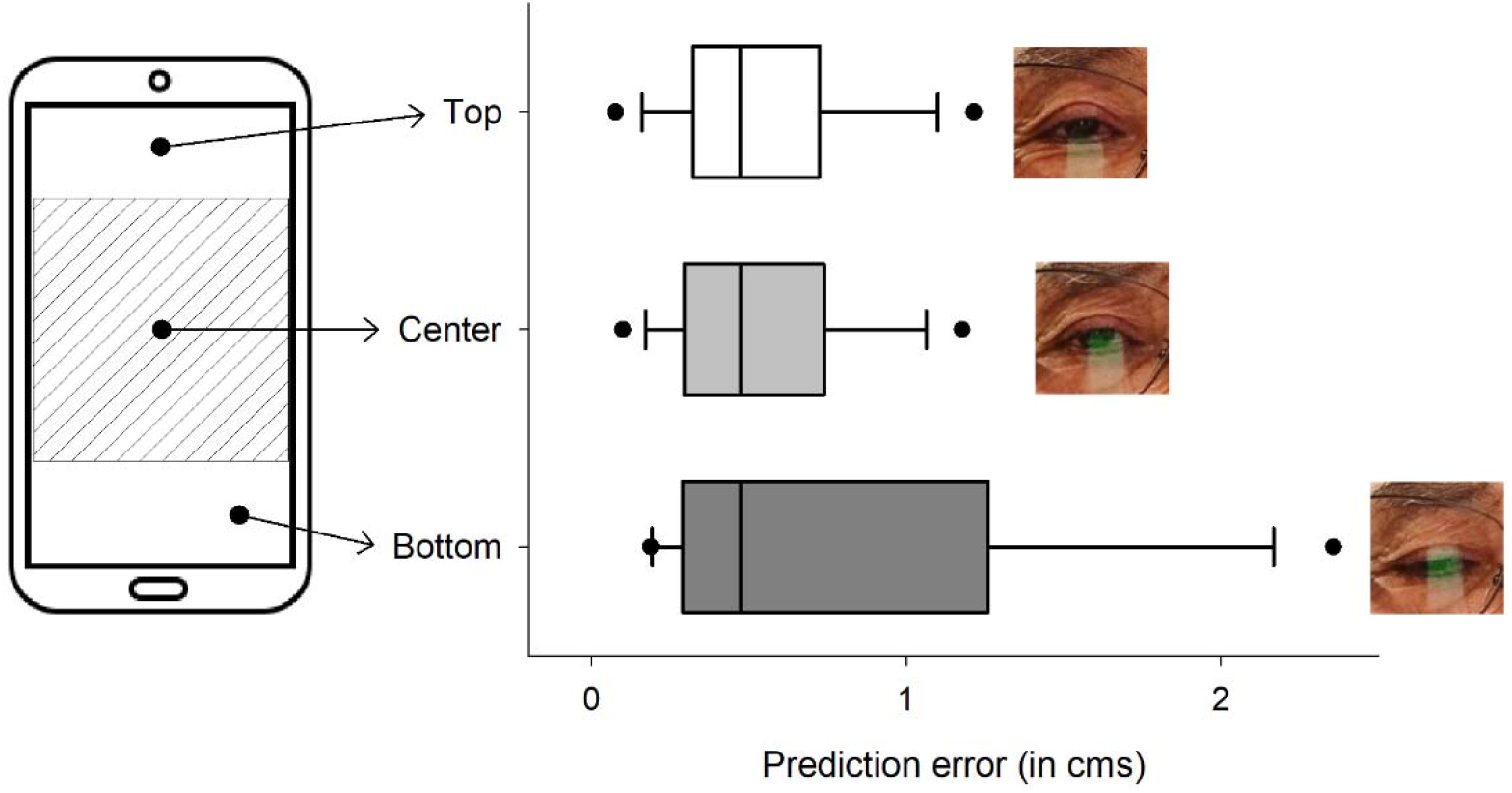
Prediction errors across the smartphone screen during calibration.

Boxplot graphs show the error rates across the smartphone screen for three regions on the smartphone screen, with outlier values representing the 5^th^ and 95^th^ percentile. Prediction errors for top, center, and bottom areas were 0.54 cm ±0.31, 0.53 cm ±0.29 and 0.79 cm ±0.69 respectively. The shaded area on the smartphone represents the approximate position of the stimuli images for the pareidolia test. As depicted in the inset images, achieving clear gaze inputs posed a challenge for the prediction models when the interpalpebral distance was at its minimum. Consequently, prediction errors variances were the highest at the bottom margins compared to top or central areas of the smartphone screen.

### Visuo-perceptual testing

Pareidolia cut-off scores are generally set at ≥2 when tested in a clinical setting^9^. 46 of the 52 participants fell within this cutoff. The remaining six participants with ≥3 pareidolias had MMSE scores above 27 and were not followed up any further. The mean response accuracies (correct answers / total answers *100) for the test were similar for both sexes (F: 97.1% ±5.9 and M: 96.0% ±9.0). Women were generally faster to respond to the stimuli with response times averaging 5.9s ±2.2 versus that with men 6.5s ±2.1. However, there was a negative trend for response accuracies and test completion time (Spearman’s rho = -0.29, *p = 0.037*).

We observed that the saliency levels did not dictate the response times (Faces vs. Noise = 6.34s ±0.6 vs 6.03s ±0.6) and did not show any significant differences across participants [t(11.5) = 1.36, p =0.21]. Gaze analysis on the dNPT showed consistent patterns for salient targets (faces) and non-targets (noise). Irrespective of the personalized eye-calibration results, heatmaps showed qualitative similarities to our prior studies on pareidolias that utilized infrared ET’s. Specifically, areas with higher saliency, such as faces, exhibited higher heat map density, indicating longer fixation and saccade densities. Welch’s independent sample t-test showed significant differences for fixations [t(11.2) = 3.75, *p =0.003*, Cohen’s d = 1.46] as well as for saccades [t(10.3) = 3.21, *p =0.009*, Cohen’s d = 1.29) for both face and noise stimuli (Figure-4).

**Figure-4:**
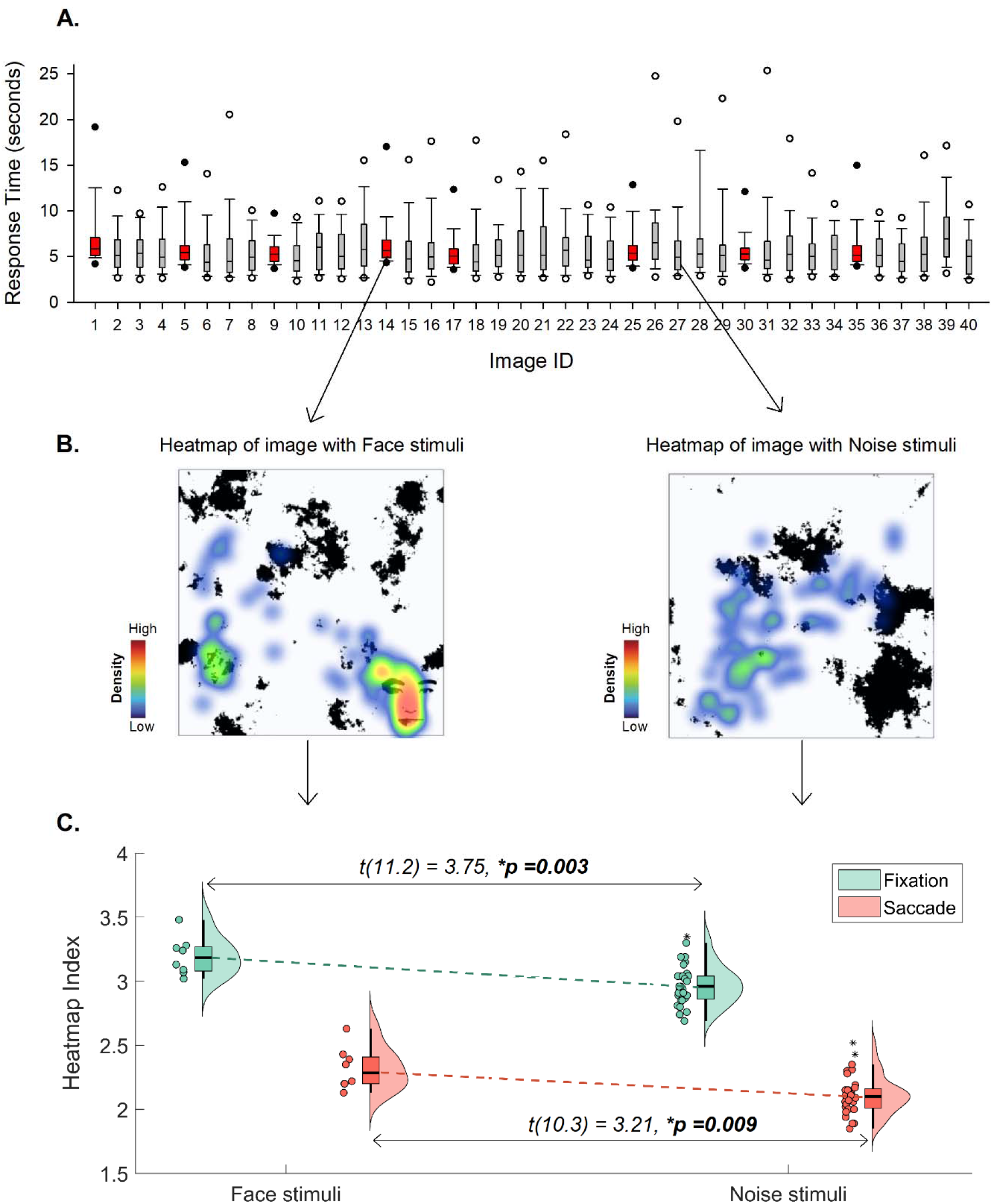
Characteristics of gaze capture across the test stimuli images. A. Shows boxplots of the response time for all 40 stimuli images with outlier values representing the 5^th^ and 95^th^ percentile. Target (Faces) stimuli are shown in red and non-targets (Noise) stimuli are shown in gray. Stimuli positions are representative according to the paper pareidolia test and for the smartphone version, they were randomized. The response time for participants was relatively similar for face and noise stimuli. B. Heatmap representation of stimuli with face image on the left and stimuli with noise on the right is shown. Warm colors (red) represent increased fixation density whereas cool colors (blue) represent search-related visual scrutiny due to saccades. C. Raincloud plots show heatmap index derived from fixation and saccades for face and noise stimuli. Fixation and saccade behavior exhibited notable differences in response to stimuli, depending on the saliency of the target.

## Discussion

Our study validated smartphone-based eye-tracking among Japanese users and demonstrated the assessment of visuo-perceptual quantification using the pareidolia test in a cohort of cognitively normal adults. We found that (i) pareidolia perception among Japanese individuals showed age-dependent variations in test completion time and response accuracy, with minimal differences based on sex, (ii) the personalized eye-tracking model showed an on-screen mean calibration error of 0.52cm when utilizing the standard (native) front-facing camera, comparable to commercial-grade eye-trackers, and (iii) gaze tracking performance on the pareidolia test exhibited distinct features of visuo-perceptual behavior linked to target saliency, which can be effectively handled using scalable smartphone technologies.

The pareidolia test is an active visual search-and-detect paradigm necessitating a knowledge-based integration of face processing^22^. The task involves relatively high cognitive demands, requiring greater attention for a preferential selection of faces^9^. As anticipated, irrespective of the sex, smartphone usage varied with age, with elderly participants exhibiting longer test completion times. Response accuracies were similar for men and women although pareidolias are suggested to be more frequent in women than in men^25^. The non-significant result could be due to the inherent nature of NPT which has reasonable class separation of signal (target) and noise (non-targets) in healthy controls^9,22^.

In exploring the pareidolia phenomenon, certain aspects were crucial in formulating the groundwork for this study. First, face perception is a stable evolutionary phenomenon although has substantial cultural differences^26–28^. Second, pareidolias are also known to vary across cultures, races, religions and are highly contingent on gender and suggestibility^15,19,20,25^. Considering these aspects to study eye-movement behavior, a general baseline ET model (e.g. MIT GazeCapture) was insufficient for our use-case. In formulating a Japanese specific model for our test, we found that our calibration accuracies substantially improved when compared to a general model. A personalized model like such could therefore be leveraged across other diverse populations.

We observed well-defined eye-movement patterns that represented attentional processes involved in top-down and bottom-up modalities of visual processing^29,30^. Because of the limited screen size, we suspect that we were unable to discern changes in the individual response times for target and non-target stimuli in our cohort. However, the differentiation between face and noise areas was clearly delineated through fixations and saccades that provide a more sensitive measure to capture attentional reorientation. In cognitively normal individuals, the saliency of faces remains robust, lowering the need to steadily augment target related information on the pareidolia test. The results evident on the heatmaps, derived from the tracking geometry, confirmed the effect of stimulus-driven fixations and saccades behavior employed for the visual search in a particular area of interest. These findings are in line with previous literature and our own research, where we utilized commercial-grade eye-trackers on both healthy individuals and patients with Parkinson’s disease^9,10,29^.

We must highlight specific control measures implemented in our experiment and acknowledge the limitations of our study. Our testing environments were carefully managed with favorable lighting conditions, and participants being tested within a very specific time of the day (early to late midday). Given that the study was conducted at a single center, it is imperative for upcoming research to examine the impact of uncontrolled lighting conditions and / or data collection from multiple centers. Approx. 60% of our users wore eyeglasses. While minimizing reflections on eyeglasses is preferable, it does not preclude our eye-tracking measurement and analysis. To an extent, it may aid in model training and generalization.

Our app software also calibrated the head / face position distance of the user automatically so that the face covered most of the camera frame. There was, however, a positional drift that was unexpectedly observed. We noted that most participants operated the smartphone with their right hand, and leaning toward a specific side may have caused a shift in the center during the test.

With respect to eye-movement calibration, during the preprocessing steps for model training, we observed a wide variance in the eyelid palpebral fissure across Japanese users in their primary gaze position. Measures to enlarge the eye-crops had no impact on the accuracy of the calibration model. Furthermore, to offset any missing frames during calibration, we performed a judicious 50-point calibration sequence which may not be ideal in real-world or clinical practice. Considering the limited screen size and that the data with missing calibration points may introduce excess noise into our inferences, we described the model accuracy only in the subset of individuals who completed the calibration fully.

As reported in a previous study^5^, even with the smartphone placed on a mounted platform, participants’ eyes may appear partially closed when they focus on the lower 20% of the screen. With a 15-minute test paradigm, it is likely that subtle shifts or adjustments while sitting upright could have impacted the head roll and pitch angles midway through the test, influencing the quality of the test results. In our cohort however, we believe this may not have affected the actual gaze tracking estimates on the pareidolia test (approx. 6 min) due to the positional coordinates of the stimuli image being at the center of the screen. Future improvements will focus on implementing a shorter 5-point or 9-point calibration sequence, correction for uncontrolled head poses while ensuring robustness and accuracy in a larger sample of participants.

### Conclusions

In summary, we demonstrated that gaze tracking quality on the pareidolia test on a smartphone is remarkable allowing us to perform complex temporal evaluations of eye-movement behavior. With their clinical relevance in several neuropsychiatric disorders^14,16,21,31^, smartphone-based pareidolia evaluation hold promise as a valuable tool for understanding cognitive processes and underlying neural mechanisms. Furthermore, our testing modality presents a scalable benefit, potentially facilitating widespread adoption and accessibility in both clinical and research settings.

## Data Availability

Data produced in the present study are available upon reasonable request to the authors

## Acknowledgements

Our sincere thanks to the individuals who participated in the study; to Megumi Matsumoto, Mika Sonoda, Xuemei Zhang from Osaka University; and to Saori Fujimoto, Takahiro Kijima, Ryoko Takei from the Dentsu Soken team (formerly ISID) for their assistance with participant recruitment and data collection.

## Author contributions

Conceptualization: GSR, EM.

Methodology: GSR, KF, EM, MS, AMA, ACS, AMJ, HM, MI, KN.

Software: GSR, AMA, ACS, VVK, AMJ, SSR, AAR.

Investigation: GSR, KF, MS, IO, YY, NY, CS, TO, DK.

Validation: GSR, KF, ACS, VVK, SSR

Formal analysis: GSR, ACS, VVK.

Resources: GSR, KF, EM, SSR, HM, MI, KN.

Data Curation: GSR, KF, AMJ, DN

Writing - Original Draft: GSR

Writing - Review & Editing: GSR, KF, EM, MS, YN, IO, YY, AMA, ACS, AMJ, SSR, AAR, NY, CS, TO, DN, HM, MI, KN.

Visualization: GSR, EM

Supervision: GSR, DN, EM, MI, HM, KN

Project administration: GSR

Funding acquisition: GSR

## Funding

G.S.R. was supported by NEDO-NEP 2024 grant; by the AMED Medical Device (AMED-SENTAN) grant, Japan, 2023-2024 (Assignment ID: J230705024); and by the KAKENHI Grant-in-Aid for Early-Career Scientists (Grant number 21K15680). The sponsors or funders did not play any role in the study design, data collection and analysis, decision to publish, or preparation of the manuscript.

## Conflict of Interest statement

The authors have no conflict of interest to report.

## Notes

### Competing Interest Statement

The authors have declared no competing interest.

### Author Declarations

The Osaka University institution review board cleared the protocol for the study to be performed in the Department of Neurology, Osaka University, Japan in accordance with the ethical standards of the Declaration of Helsinki (IRB Approval number 22307).

## References

1. Eckstein, M. K., Guerra-Carrillo, B., Miller Singley, A. T. & Bunge, S. A. Beyond eye gaze: What else can eyetracking reveal about cognition and cognitive development? Developmental Cognitive Neuroscience 25, 69–91 (2017).

2. Mele, M. L. & Federici, S. Gaze and eye-tracking solutions for psychological research. Cogn Process 13, 261–265 (2012).

3. Anderson, T. J. & MacAskill, M. R. Eye movements in patients with neurodegenerative disorders. Nat Rev Neurol 9, 74–85 (2013).

4. Zimmermann, J., Vazquez, Y., Glimcher, P. W., Pesaran, B. & Louie, K. *Oculomatic*: High speed, reliable, and accurate open-source eye tracking for humans and non-human primates. Journal of Neuroscience Methods 270, 138–146 (2016).

5. Valliappan, N. et al. Accelerating eye movement research via accurate and affordable smartphone eye tracking. Nat Commun 11, 4553 (2020).

6. Zhang, X., Sugano, Y., Fritz, M. & Bulling, A. Appearance-Based Gaze Estimation in the Wild. in 2015 IEEE Conference on Computer Vision and Pattern Recognition (CVPR) 4511– 4520 (2015). doi:10.1109/CVPR.2015.7299081.

7. Krafka, K. et al. Eye Tracking for Everyone. MIT Web Domain (2016).

8. Pfeiffer, U. J., Vogeley, K. & Schilbach, L. From gaze cueing to dual eye-tracking: Novel approaches to investigate the neural correlates of gaze in social interaction. Neuroscience & Biobehavioral Reviews 37, 2516–2528 (2013).

9. Revankar, G. S. et al. Ocular fixations and presaccadic potentials to explain pareidolias in Parkinson’s disease. Brain Commun 2, fcaa073 (2020).

10. Liu, J. et al. Seeing Jesus in toast: Neural and behavioral correlates of face pareidolia. Cortex 53, 60–77 (2014).

11. Revankar, G. et al. Pre-stimulus low-alpha frontal networks are associated with pareidolias in Parkinson’s disease. Brain Connectivity (2021) doi:10.1089/brain.2020.0992.

12. Linszen, M. M. J. et al. Understanding hallucinations in probable Alzheimer’s disease: Very low prevalence rates in a tertiary memory clinic. *Alzheimer’s & Dementia: Diagnosis*, Assessment & Disease Monitoring 10, 358–362 (2018).

13. Hamilton, C. A. et al. Utility of the pareidolia test in mild cognitive impairment with Lewy bodies and Alzheimer’s disease. Int J Geriatr Psychiatry 36, 1407–1414 (2021).

14. Uchiyama, M. et al. Pareidolias: complex visual illusions in dementia with Lewy bodies. Brain 135, 2458–2469 (2012).

15. Yokoi, K. et al. Hallucinators find meaning in noises: pareidolic illusions in dementia with Lewy bodies. Neuropsychologia 56, 245–254 (2014).

16. Nishio, Y. et al. Deconstructing psychosis and misperception symptoms in Parkinson’s disease. *Journal of Neurology*, Neurosurgery & Psychiatry 88, 722–729 (2017).

17. O’Brien, J. et al. Visual hallucinations in neurological and ophthalmological disease: pathophysiology and management. *Journal of Neurology*, Neurosurgery & Psychiatry 91, 512–519 (2020).

18. Weil, R. S. et al. Visual dysfunction in Parkinson’s disease. Brain 139, 2827–2843 (2016).

19. Romagnano, V., Sokolov, A. N., Fallgatter, A. J. & Pavlova, M. A. Do subtle cultural differences sculpt face pareidolia? Schizophr 9, 1–7 (2023).

20. Wang, C., Yu, L., Mo, Y., Wood, L. C. & Goon, C. Pareidolia in a Built Environment as a Complex Phenomenological Ambiguous Stimuli. Int J Environ Res Public Health 19, 5163 (2022).

21. Revankar, G. S. et al. Perceptual constancy of pareidolias across paper and digital testing formats in neurodegenerative diseases. 2024.02.08.24302504 Preprint at 10.1101/2024.02.08.24302504 (2024).

22. Mamiya, Y. et al. The Pareidolia Test: A Simple Neuropsychological Test Measuring Visual Hallucination-Like Illusions. PLoS ONE 11, e0154713 (2016).

23. Ideno, Y., Takayama, M., Hayashi, K., Takagi, H. & Sugai, Y. Evaluation of a Japanese version of the Mini-Mental State Examination in elderly persons. Geriatrics & Gerontology International 12, 310–316 (2012).

24. Watanabe, H. et al. Negative mood invites psychotic false perception in dementia. PLoS One 13, (2018).

25. Proverbio, A. M. & Galli, J. Women are better at seeing faces where there are none: an ERP study of face pareidolia. Soc Cogn Affect Neurosci 11, 1501–1512 (2016).

26. Little, A. C., Jones, B. C. & DeBruine, L. M. Facial attractiveness: evolutionary based research. Philos Trans R Soc Lond B Biol Sci 366, 1638–1659 (2011).

27. Kelly, D. J. et al. Developing cultural differences in face processing. Developmental Science 14, 1176–1184 (2011).

28. Oruc, I., Balas, B. & Landy, M. S. Face perception: A brief journey through recent discoveries and current directions. Vision Research 157, 1–9 (2019).

29. Fischer, T., Graupner, S.-T., Velichkovsky, B. M. & Pannasch, S. Attentional dynamics during free picture viewing: Evidence from oculomotor behavior and electrocortical activity. Front Syst Neurosci 7, 17 (2013).

30. Connor, C. E., Egeth, H. E. & Yantis, S. Visual Attention: Bottom-Up Versus Top-Down. Current Biology 14, R850–R852 (2004).

31. Abo Hamza, E. G., Kéri, S., Csigó, K., Bedewy, D. & Moustafa, A. A. Pareidolia in Schizophrenia and Bipolar Disorder. Front Psychiatry 12, 746734 (2021).

