## Supplementary material for "Mapping smartphone-based eye-tracking behavior across Japanese individuals on the pareidolia test": Suppl data

**Supplementary Figure-1. Experimental setup and characteristics of eye crop data from the cohort.**

****Contact corresponding author for image****

A. The experiment setup shows a representative user (consent provided for the image), visualizing images on the smartphone mounted on a stand. In this controlled setting, the front-camera lined up at the participant eye-brow level when performing the test with an approx. distance of 30cm between the user and the smartphone. The app software provided feedback on the user’s head position ensuring head angles at an appropriate range for testing. Head roll and pitch angle range were set between 0 to 15 degrees and 71 to 83 degrees respectively. Head yaw angle was not considered since it did not affect the data collection or model output.
B. Representative examples of eyelid interpalpebral fissure sizes across 6 different participants on the central frame. On close observation, the reflection of the smartphone screen is noted in participants with eyeglasses. However, our model accuracy remained stable irrespective of the use of eyeglasses.

**Supplementary Figure-2. Block diagram of the eye-tracking model architecture.**

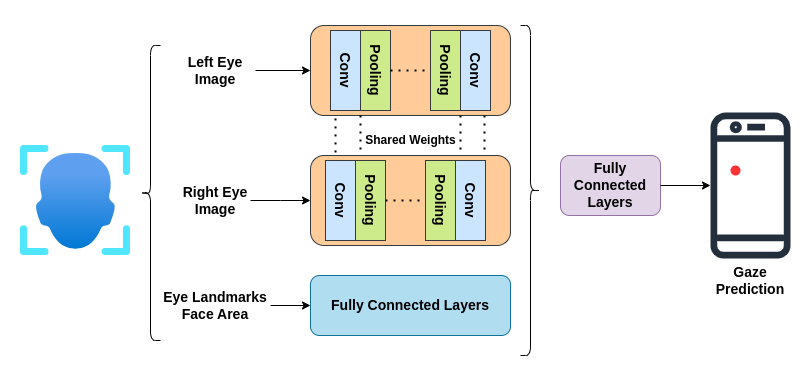

Shows the base-model architecture trained on MIT GazeCapture and Japanese datasets. We refined the architecture from the one described by Valliappan et.al.^5^. Eye regions were cropped based on the eye corner landmarks, scaled to 128 × 128 × 3 pixels, and fed through two identical convolutional layer blocks. Other features like eye corners and face area were fed through fully connected layers blocks to ultimately combine and get the gaze location on screen. We used average pooling and ReLU activation functions in the model.

**Supplementary Figure-3. Calibration grid and gaze prediction**

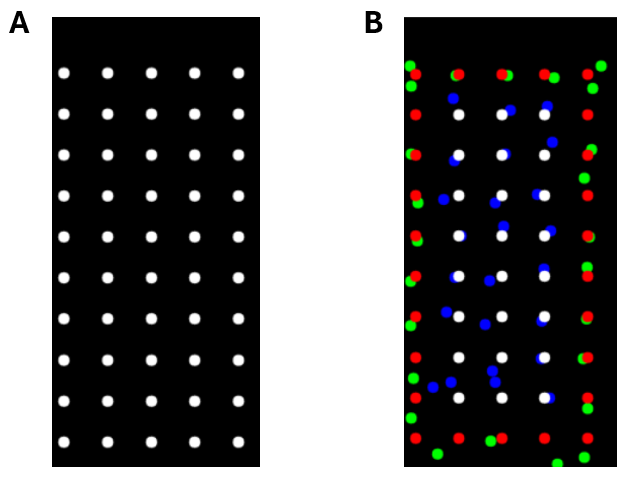

Figure shows a representative example of a calibration sequence for a single participant. (A) A 50-dot calibration grid was used with each dot displayed serially for 3 seconds with a total calibration duration of 150 seconds. (B) The red / white dots are ground truth, and green / blue dots are predictions of gaze location shown for margins and central areas. Prediction errors were lower at the center of the screen and the highest errors were observed around the corners and margins of the screen.
